# AI-based Speech Error Detection to Differentiate Primary Progressive Aphasia Variants

**DOI:** 10.64898/2026.02.23.26346899

**Authors:** Jet M.J. Vonk, Jiachen Lian, Cheol J. Cho, Giada Antonicelli, Zoe Ezzes, Lisa D. Wauters, Willa Keegan-Rodewald, G. Lynn Kurteff, Diana Alejandra Rodriguez, Nina Dronkers, Maya L. Henry, Zachary A. Miller, Maria Luisa Mandelli, Gopala K. Anumanchipalli, Maria Luisa Gorno-Tempini

## Abstract

**Background:** Artificial Intelligence (AI) based approaches to speech analysis have the potential to assist with objective speech error analysis in aphasia but off-the shelf tools often fail to detect speech errors due to prioritizing “fluent transcription.” Speech production errors (dysfluencies) are hallmark diagnostic features of the nonfluent (nfvPPA) and logopenic (lvPPA) variants of primary progressive aphasia, yet they can be challenging to detect and characterize even by expert clinicians. This study aimed to evaluate whether the novel automated lightweight Scalable Speech Dysfluency Modeling system (SSDM-L), specifically designed to detect dysfluencies, could accurately distinguish PPA variants using voice recordings of individuals reading a brief passage.

**Method:** Participants included a total of 104 individuals, 40 with nfvPPA, 40 with lvPPA (matched on disease severity), and 24 healthy controls who read aloud the ‘Grandfather Passage’ as part of a widely used motor speech evaluation (MSE). We automatically extracted ten speech error (dysfluency) variables using SSDM-L, including insertions, replacements, and deletions at both phoneme- and word-levels, and phoneme-level prolongations and repetitions. Group differences were assessed via ANCOVAs controlling for age, education, and disease severity (MMSE, CDR sum-of-boxes). To test clinical relevance, we performed correlation analyses with MSE ratings provided by experienced speech-language pathologists (i.e., gold standard) within the nfvPPA group. Classification performance was assessed by training random forest and XGBoost machine-learning models including 5-fold cross-validation.

**Results:** All individuals read the entire passage in less than five minutes. SSDM-L detected eight of the ten predefined dysfluency features at sufficient frequency to include them in subsequent analyses. All eight features distinguished PPA from controls (p<.006). Individuals with nfvPPA made more errors than the lvPPA group on every feature (all p<.023). Each feature showed a moderate positive correlation with a global MSE apraxia/dysarthria score (r=.31–.56; p<.001-.053). Together, the eight features were able to classify nfvPPA versus lvPPA at AUC=.806 (random forest) and AUC=.776 (XGBoost).

**Discussion:** AI-based automated speech error analysis accurately distinguished nfvPPA and lvPPA variants using a brief reading task. This quick error-sensitive scalable AI system has the potential of providing a practical tool to aid diagnosis in aphasia and motor speech disorders.

## Introduction

Advances in speech technologies, such as OpenAI Whisper, Google Speech-to-Text, and others, have significantly improved transcription through automated speech recognition (ASR) and subsequent speech analysis of large-scale spoken language datasets.^1,2^ By providing automated transcriptions, these tools facilitate extracting linguistic features such as lexical diversity and syntactic complexity in individuals who produce relatively fluent speech and have shown promise as cognitive markers in dementias such as Alzheimer’s disease.^3–5^ However, applying these technologies to the dysfluent speech of individuals who present with specific language and articulation difficulties is far more challenging. Speech produced by such individuals often contains word- and phoneme-level errors (dysfluencies) that are critical for diagnostic purposes but are poorly captured by standard ASR systems, creating limitations in the use of current tools for these populations.^6^ Off-the-shelf ASR systems such as Whisper are biased toward fluent speech transcription and often “correct away” clinically meaningful speech errors like substitutions, repetitions, deletions, and pauses.^7^ These dysfluencies are essential for early detection and monitoring of conditions such as primary progressive aphasia (PPA), a neurodegenerative disease that selectively affects speech and language functions.^8^

Among the three canonical variants of PPA, the nonfluent/agrammatic variant (nfvPPA), with motor speech and grammatical deficits,^9^ and the logopenic variant (lvPPA), with phonological deficits,^10,11^ are the two that show clear dysfluencies in spontaneous speech. Distinguishing nfvPPA and lvPPA is particularly challenging,^12^ as nfvPPA involves apraxia of speech and phonetic errors (i.e., focused on how a sound is produced), while lvPPA produces phonemic errors (i.e., focused on what sound is intended).^13,14^ Distinguishing between these errors requires specialized expertise from speech-language pathologists (SLPs), and, even then, inter-rater reliability among SLPs often varies.^15^ Advanced AI techniques tailored to detect a range of error patterns within speech samples across patients could improve diagnostic accuracy.^16,17^ Differential diagnosis between these two variants is crucial since lvPPA is most often caused by Alzheimer’s disease pathology,^11,18,19^ while nvPPA is related to frontotemporal lobar degeneration,^19^ each with distinct implications for prognosis, treatment, clinical trial eligibility, and disease monitoring.

To address this limitation, we developed a lightweight Scalable Speech Dysfluency Modeling system (SSDM-L), a novel AI framework designed to accurately transcribe and analyze speech errors.^20^ This system was created to effectively characterize speech errors within a given speech sample against a known ground-truth target script. SSDM-L is explainable and specifically designed to be sensitive to speech dysfluencies. SSDM-L comprises three sub-modules: a) Dysfluent-WFST44: a high-performance phonetic decoder that transcribes the phonetic sequence of the patient’s speech sample;^21^ b) Neural-LCS45: an alignment module that maps the phonetic sequence from (a) to the reference phoneme sequence (e.g., the Grandfather script);^22^ and c) Speech Error Analysis Module: a component that uses the alignment produced in (b) to categorize specific types of speech errors.^20^ SSDM-L is capable of detecting ten distinct types of dysfluencies, including insertions, repetitions, deletions, and replacements at both phoneme and word levels, thus capturing a wide range of error patterns underlying diverse speech-language disorders.

This study evaluated whether the SSDM-L AI system could reliably capture speech production errors in a large cohort of well-characterized individuals with nfvPPA and lvPPA. We investigated whether these dysfluencies obtained from voice recording of patients with PPA reading a short 2-minute paragraph would: (1) identify group-level differences between PPA variants; (2) correlate with expert SLP severity ratings for motor speech impairment as a proxy for speech output deficits; and (3) accurately classify individuals into nfvPPA and lvPPA. We hypothesized that dysfluencies identified by SSDM-L would be more prevalent in nfvPPA than in lvPPA, reflecting the fluency deficits typical of the nfvPPA variant. The overall goal of this study is to demonstrate that advanced, automated AI speech modeling techniques hold the potential to aid and scale diagnostic precision of progressive speech and language disorders.

## Method

### Participants

This study included 104 participants: 40 diagnosed with nonfluent/agrammatic variant PPA (nfvPPA), 40 with logopenic variant PPA (lvPPA), and 24 cognitively healthy controls. All participants were evaluated at the University of California, San Francisco (UCSF) Edward and Pearl Fein Memory and Aging Center (MAC) PPA programs. Individuals seen at the MAC undergo detailed clinical, neuropsychological, and imaging assessments. Healthy controls were recruited primarily through the Brain Aging Network for Cognitive Health (BrANCH) cohort and were cognitively assessed to confirm the absence of neurological or psychiatric disorders.

We identified all cases in the UCSF Fein MAC PPA program database with a clinical diagnosis of nfvPPA or lvPPA, following established diagnostic criteria,^8^ who had completed research visits since 2001, had an available recorded Grandfather Passage reading task,^23,24^ and were native English speakers. To focus on early-stage differentiation between PPA variants, we excluded individuals who did not complete at least nearly half of the passage (4/9 sentences, 45% of words), had Mini-Mental State Examination (MMSE) scores below 17, or had Clinical Dementia Rating (CDR) sum of boxes scores greater than 9. These thresholds were selected to exclude individuals with moderate or severe dementia, generally considered as CDR sum of boxes>9 and MMSE<18. We used a slightly lower cut-off on MMSE to account for the language-dependent nature of the MMSE, which often underestimates cognitive function in PPA.^25^ Data from each participant’s earliest available research visit that included administration of the Grandfather Passage were used for analysis. To minimize confounding factors and account for differences in PPA subgroup sizes, we performed frequency matching to match PPA groups on age, sex/gender, education, MMSE, CDR, and group size (all p>.05), while also ensuring the control group did not differ from either PPA group on age, sex/gender, and education (all p>.05). A detailed speech and language evaluation is completed with every individual participant as previously described.^26^ All participants or caregivers provided informed consent following procedures aligned with the Declaration of Helsinki, and the study was approved by the UCSF Institutional Review Board (IRB # 10-03946).

### Neuroimaging analysis of group-level atrophy patterns

A group-level neuroimaging analysis was conducted to confirm typical patterns of atrophy for each variant. MRI T1 images were acquired for 92 subjects (39 nfvPPA, 38 lvPPA, 15 controls) with sequences, previously described, on either 1.5T (n=7),^26^ 3T (n=81),^27^ or 4T (n=4).^28^ After a visual quality check, one participant from the nfvPPA group was excluded for excessive motion. All the structural images underwent standard preprocessing for thickness analysis using the Computational Anatomy Toolbox 12 (CAT12) version 12.9 (<u>dbm.neuro.uni-jena.de/cat</u>) in Statistical Parametric Mapping 12 software version 7771 (<u>fil.ion.ucl.ac.uk/spm/software/spm12</u>) as detailed in Mandelli et al. (2023).^29^

Whole-brain analyses of between-group differences in cortical thickness (nfvPPA vs. controls; lvPPA vs. controls) were estimated using independent sample t-tests covaried for age and sex. Alpha levels were set at a peak-level threshold of p<.05 family-wise error (FWE) corrected. Visualization of group atrophy patterns was performed using CAT12.

### Motor Speech Evaluation including the Grandfather Passage

Each participant underwent a Motor Speech Evaluation (MSE)^30^ during their initial clinical language assessment, which was recorded on video. As part of this evaluation, participants read the Grandfather Passage aloud; the audio from this recording was extracted for use in the present study’s speech analysis.

The MSE is a clinical assessment designed to detect and diagnose motor speech disorders, specifically AOS and dysarthria,^31^ and is widely used in clinical research and diagnosis, including in individuals with PPA.^9,13,32^ The test protocol consists of nine subtests that evaluate various aspects of speech production, including automatic speech production (i.e., counting from 1-20), sustained phonation, sequential motion rates (i.e., repeating /pʌpʌpʌ/; /tʌtʌtʌ/; /kʌkʌkʌ/), alternating motion rates (i.e., repeating /pʌtʌkʌ/), multisyllabic word repetition, production of words of increasing length, sentence repetition, and reading aloud of the Grandfather Passage. Each participant receives a 0-7 severity rating for both AOS and dysarthria, which is informed by characteristic features observed across the tasks (0: Within normal limits, 1: Slight, 2: Mild, 3: Mild to moderate, 4: Moderate, 5: Moderate to severe, 6: Severe, 7: Profound). Notably, the MSE is designed to detect motor speech and not phonological errors, thus SLPs are likely to only rate errors that are considered motoric in nature.

The Grandfather Passage is a brief, phonetically balanced paragraph used to elicit connected speech in a controlled context through reading aloud. Originally developed by Charles Van Riper in 1963^23^ and adapted by Darley, Aronson, & Brown (1975)^24^ for use in speech and language assessment, the passage includes all phonemes in the English language and contains complex phonotactic combinations (e.g., *frock*, *zest*), as well as syntactic and semantic variety.^33^ These characteristics make it particularly useful for eliciting speech production deficits. The brevity of the passage (typically 1-3 minutes) makes it efficient to administer while still providing a rich sample for analysis. Because the spoken text is predetermined, the Grandfather Passage provides an optimal context for validating the SSDM-L technique using phoneme-level forced alignment.

### AI-based automated speech transcription and feature extraction

SSDM-L comprises a phoneme transcription module, phoneme alignment module, phoneme-level dysfluency detector, as well as word-level transcription, alignment, and dysfluency detection modules. The phoneme transcription component captures the actual produced phonemes, adapted from Dysfluent-WFST,^21^ a state-of-the-art phonetic transcriber. With the Grandfather Passage serving as the reference text, SSDM-L applies the Neural Longest Common Subsequence (NLCS)^22^ algorithm to align the uttered speech with the reference text at the phoneme level. Phonetic dysfluencies are detected as follows: if the uttered phoneme matches the reference phoneme, no dysfluency is reported; otherwise, insertions, omissions, substitutions, or repetitions are identified. Each dysfluency is reported at most once for each reference phoneme. An analogous set of four features is extracted at the word level. Additionally, SSDM-L integrates Unconstrained Dysfluency Modeling (UDM),^34^ a state-of-the-art dysfluent phoneme aligner, to detect irregular phoneme durations, particularly for vowels. If the duration exceeds a predefined threshold, phoneme prolongation is reported. UDM also detects irregular intra-word pauses, which are counted at the word level. Thus, SSDM-L extracts raw counts of ten features in total (Table 1): phoneme insertion, phoneme replacement, phoneme repetition, phoneme deletion, phoneme prolongation, word insertion, word replacement, word repetition, word deletion, and intra-word pauses (i.e., pauses within a word). Of note, none of these features are consistently or accurately captured by off-the-shelf ASR tools. We computed normalized values for each feature by dividing the dysfluency counts by the total number of corresponding phonemes or words, respectively.

**Table 1.**
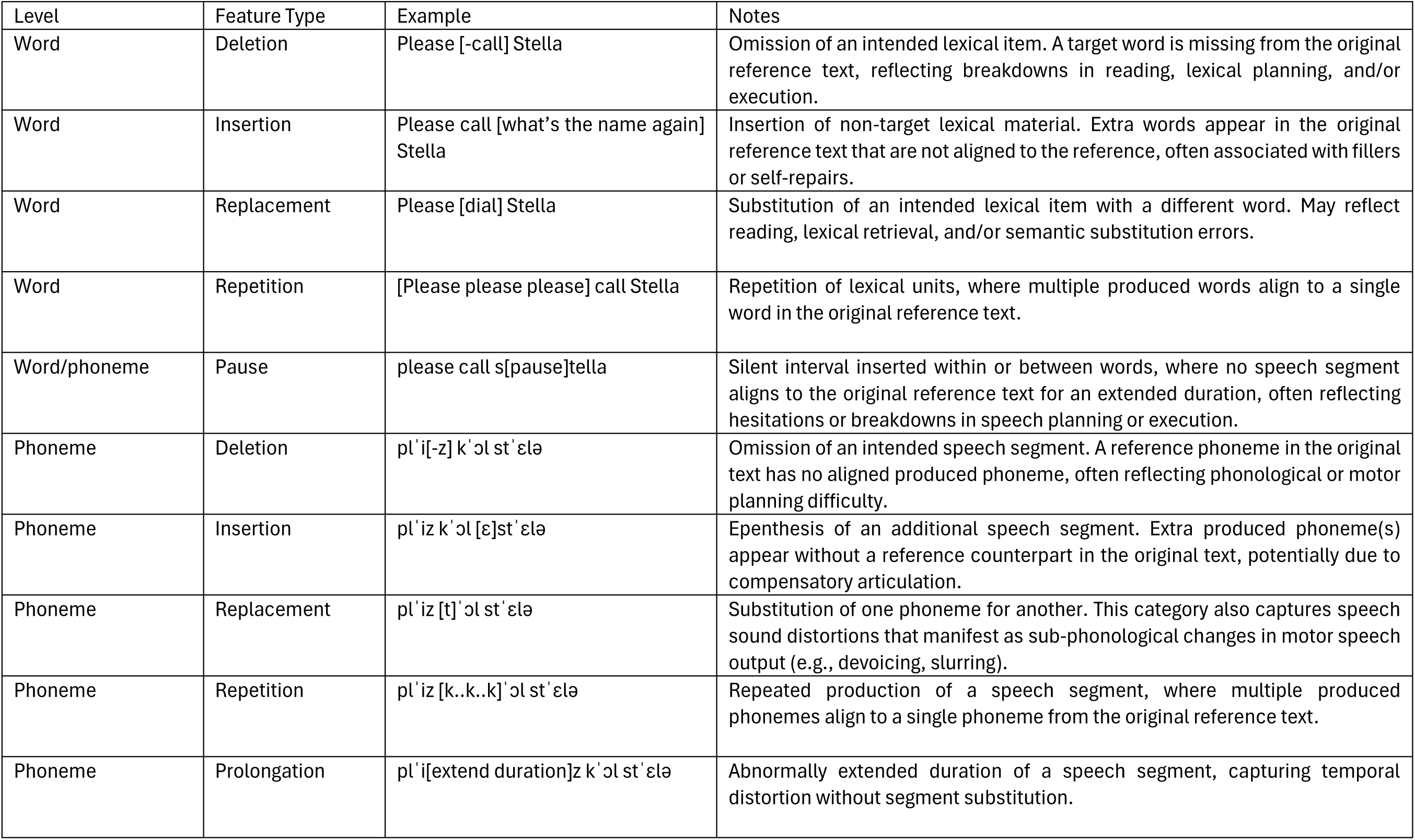
Features extracted by SSDM-L with examples.

### Statistical analysis

Sample characteristics and distributions of extracted speech features were examined using descriptive statistics, general linear models, chi-square tests, and Pearson correlation coefficients. Because participants did not always complete the full standardized passage, we normalized phoneme- and word-level error counts by dividing each by the total number of phonemes or words read, respectively. Group differences in individual features were assessed using analysis of variance (ANOVA; unadjusted) and analysis of covariance (ANCOVA), adjusting for age, education, and disease severity (z-scored composite of CDR Sum of Boxes and MMSE^35^). Post-hoc pairwise comparisons between diagnostic groups were corrected for multiple testing using the false discovery rate (FDR).

To assess whether the AI-extracted dysfluencies correlate with expert SLP ratings of motor speech output difficulties, we examined their relationship with clinical ratings of the overall MSE AOS and dysarthria ratings in the nfvPPA group. Pearson correlation coefficients were computed between each feature and the clinician-rated severity scores for AOS and dysarthria, as well as a composite “global” MSE score defined as the sum of the AOS and dysarthria ratings (higher scores indicating more severe impairment). While AOS and dysarthria are different motor speech disorders with different neurological substrates (and the MSE ratings would usually not be combined by a practicing SLP), we aim at correlating the AI-based scores with a general, SLP-derived score reflecting the overall severity of motor speech output errors that patients make, regardless of their pathophysiological origin. In other words, the purpose of our global MSE score is not to make claims about the substrates of AOS and dysarthria, but to serve as a proxy of overall severity of motor speech impairment that is easily comparable to the SSDM-L system’s estimates of severity.

To evaluate the utility of the extracted features in distinguishing between diagnostic groups and to ensure robustness across modeling approaches, we trained two machine learning classifiers: random forest and extreme gradient boosting (XGBoost). Random forest was included as a widely adopted, interpretable machine learning method that reduces overfitting through ensemble learning and has demonstrated strong performance in prior classification studies using automated speech data in dementia research.^36^ XGBoost was selected for its ability to handle small sample sizes, deliver stable performance with minimal tuning, and its similarly proven success in classification tasks involving speech-based markers of neurodegenerative disease.^37,38^ The data were split into 65% training and 35% testing sets, with model performance assessed using five-fold cross-validation within the training set. Area under the receiver operating characteristic curve (AUC) was used as an evaluation metric. All extracted speech error (dysfluency) features used in the ANCOVA analysis were included in the classification models without prior feature selection. All analyses and visualizations were conducted in R (version 4.4.2) using a range of packages as listed in the analysis code, which is available at https://github.com/jmjvonk.

### Data availability

UCSF Fein MAC ethics approval does not permit depositing anonymized study data in a public repository. Data can be obtained on reasonable request via the UCSF Fein MAC Resource Request form (http://memory.ucsf.edu/resources/data). Access is granted to named investigators under ethical regulations governing reuse of sensitive data, and all requests require a Material Transfer Agreement (MTA) processed through UCSF (see https://icd.ucsf.edu/material-transfer-and-data-agreements). Requests for commercial use will not be approved.

## Results

### Participant and feature characteristics

Sample characteristics for demographics, disease severity, standard cognitive and speech-language tests, and MSE scores across groups are summarized in Table 2. As the cognitively healthy control, lvPPA, and nfvPPA groups were matched on age, sex/gender, and education, there were no significant differences among groups for these variables (all p>.05). There were also no differences between groups on race/ethnicity or handedness (all p>.05). The cognitively healthy control group had higher MMSE scores and lower CDR Sum of Boxes scores than both PPA groups, while the lvPPA and nfvPPA groups were matched on these variables and thus did not differ from each other in disease severity scores. The two PPA variants showed speech and language characteristics compatible with their diagnosis, with lvPPA showing greater repetition and confrontation naming deficits, and the nfvPPA showing greater fluency and motor speech difficulties.^8^ The lvPPA group scored 0 on MSE severity ratings for both AOS and dysarthria, except for one individual with an AOS score of 1, compatible with the MSE’s focus on motor speech errors, whereby other types of speech production errors are not captured. In the nfvPPA group, all individuals had at least one score higher than 0 on either AOS or dysarthria ratings; 5/40 had dysarthria without AOS, 9/40 had AOS without dysarthria, and 26/40 had co-occurring AOS and dysarthria. For 16/40 AOS was more severe (>1 difference in score) than dysarthria, for 9/40 dysarthria was more severe (>1 difference in score) than AOS, and 15/40 had relatively equal scores (max 1 difference in score) on AOS and dysarthria. Scores on the AOS (max 4) and dysarthria (max 5) ratings in the nfvPPA group were in the mild range,^13^ consistent with the early disease stage of the selected individuals.

**Table 2.**
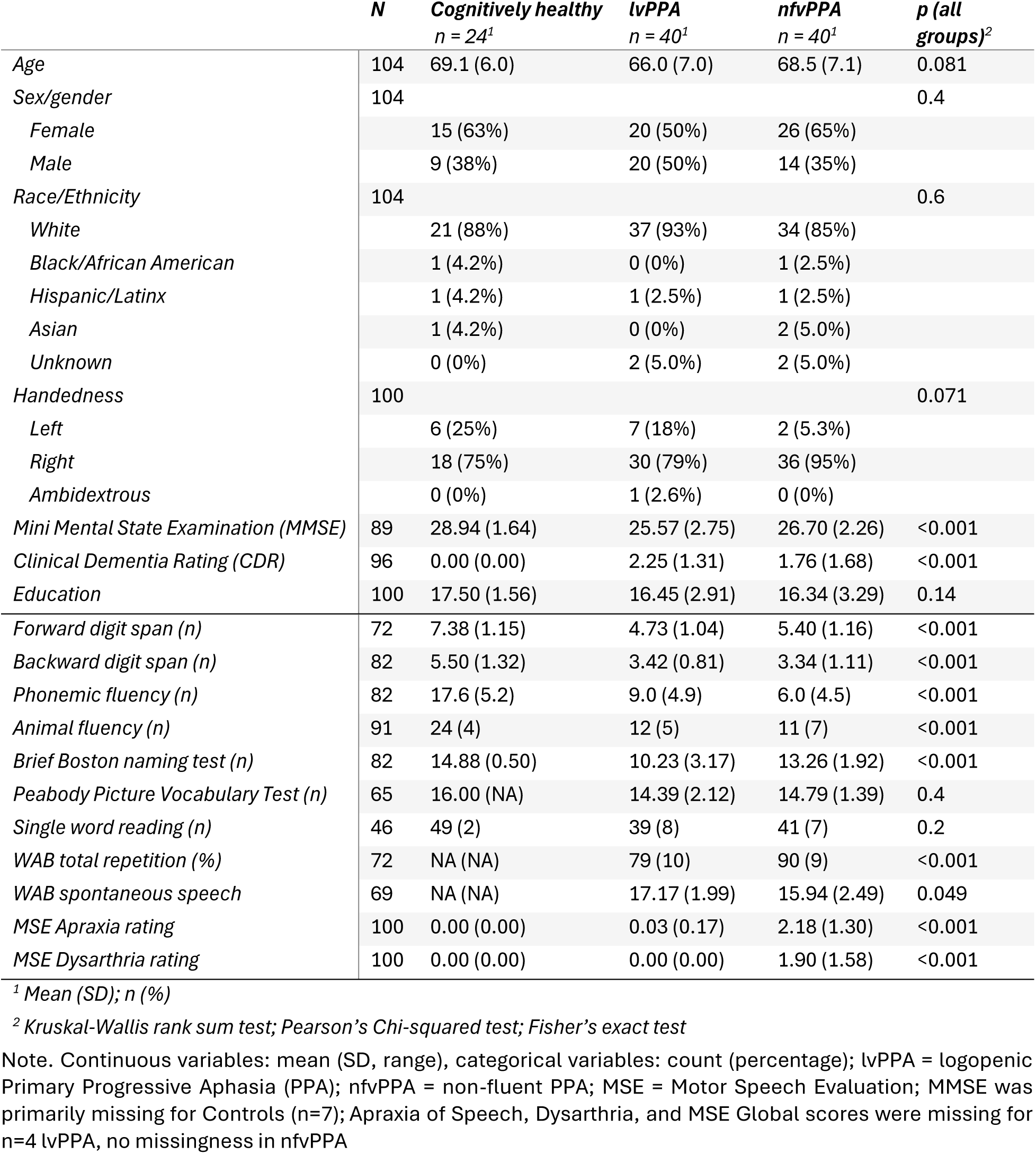
Participant characteristics.

The average time to complete reading the Grandfather Passage aloud was 48.8 seconds (SD = 5.7, range = 34.6-60.8) for cognitively healthy participants, 76.6 seconds (SD = 30.4, range = 41.5-168.6) for the lvPPA group, and 107.0 seconds (SD = 48.8, range = 51.5-285.0) for the nfvPPA group. SSDM-L automatically extracted counts of ten phoneme-level and word-level speech errors during reading aloud the Grandfather Passage: phoneme insertion, phoneme replacement, phoneme repetition, phoneme deletion, phoneme prolongation, word insertion, word replacement, word repetition, word deletion, and intra-word pauses. Inspection of feature distributions across all participants showed that phoneme-level errors were common, each occurring 94-104 times. Word-level features were on average slightly less frequent but still widely present, with word insertion occurring 93 times, word replacement 97 times, and word deletion 69 times. In contrast, word repetition and intra-word pauses were rare, occurring only 18 and 24 times, respectively, across all participants. Because word repetition and intra-word pause errors were infrequently captured by the SSDM-L system, they were excluded from further group comparisons and classification analyses due to low prevalence.

### Neuroimaging atrophy patterns

Participants in the nfvPPA group exhibited left-sided atrophy in the inferior frontal gyrus, supplementary motor area, and precentral gyrus (Figure 1A). Participants in the lvPPA group exhibited left-sided atrophy in the temporal-parietal junction and temporal lobe, particularly the superior, middle, and inferior temporal gyri (Figure 1B).

**Figure 1.**
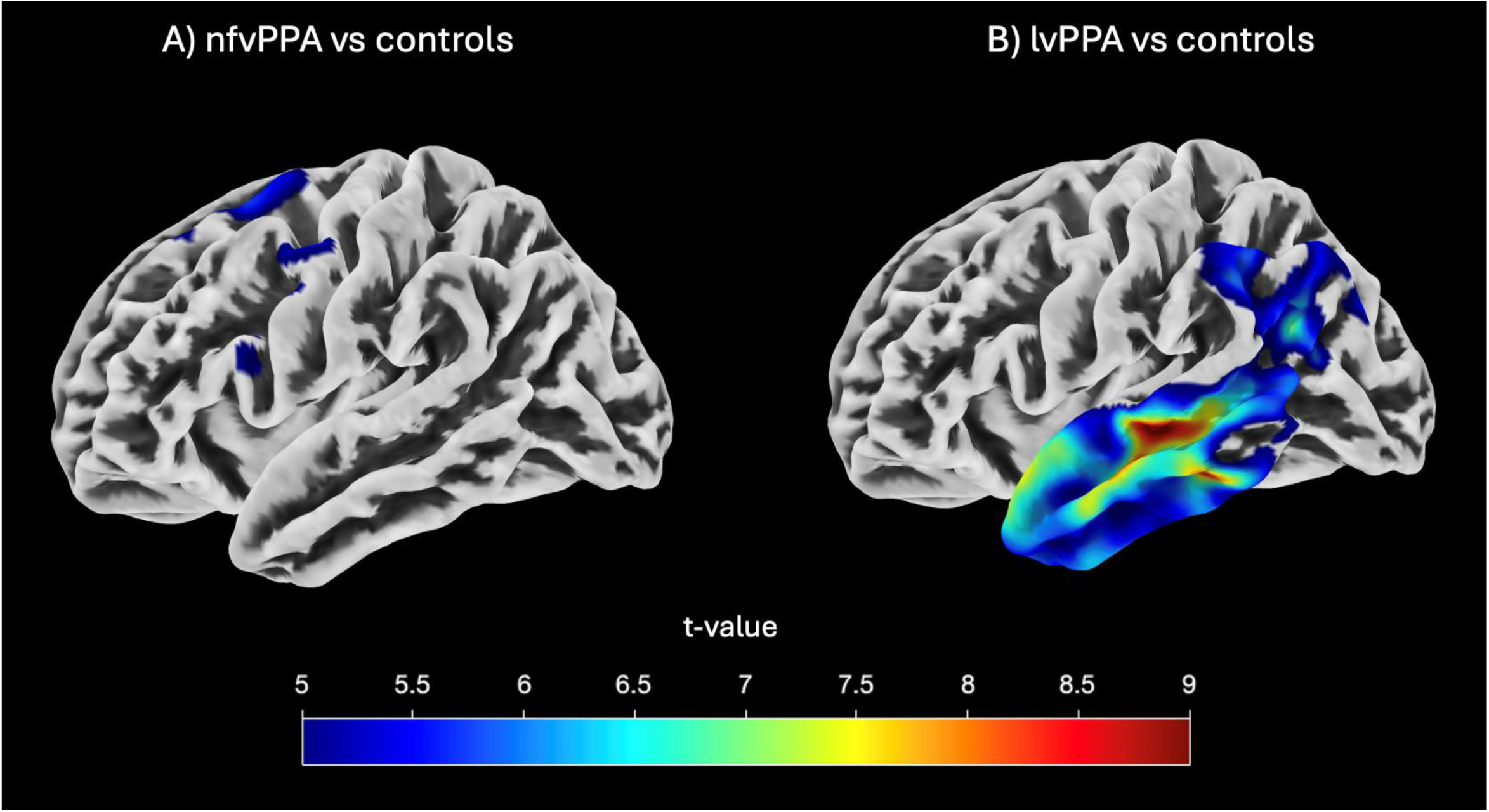
Brain atrophy patterns

### Group differences

Differences in error counts on the eight remaining phoneme- and word-level features between diagnostic groups is visualized in the boxplots in Figure 2. Each feature shows a similar visual pattern, in which the nfvPPA group made the most errors, followed by the lvPPA group, followed by almost no errors in the control group. AN(C)OVA models to statistically assess group differences in features showed similar results unadjusted or adjusted for covariates (i.e., age, education, disease severity); this report focuses primarily on the adjusted models. All features differed across the three diagnostic groups (all p<.006). Pairwise comparisons (FDR-corrected) showed the nfvPPA group made more errors than the lvPPA group on every feature (all p<.023; Table 3). While unadjusted, the lvPPA group made significantly more errors than the control group on nearly every feature except word deletion (Fig 2); no group differences between lvPPA and controls remained significant after adjustment for covariates and FDR correction in the pairwise comparisons (all p>.084). In contrast, the nfvPPA group robustly made more errors than the control group on all features in unadjusted and adjusted models, with all p<.024 after adjustment for covariates and FDR correction, except for word deletion at p=.055.

**Figure 2.**
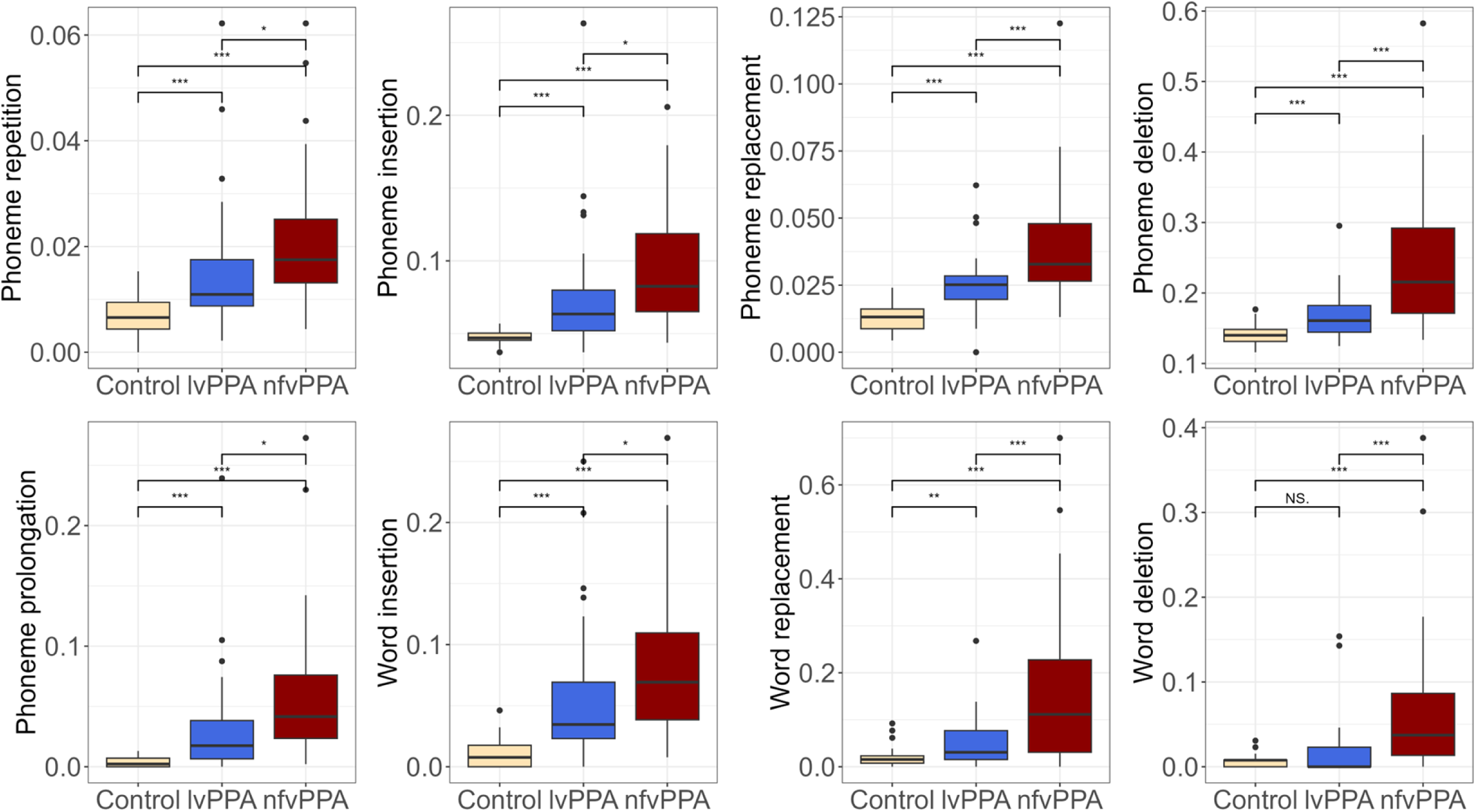
Normalized^†^ counts of phoneme- and word-level errors across diagnostic groups (Control n=24; lvPPA n=40; nfvPPA n=40) Note. ^†^Scores were normalized by dividing phoneme- and word-level error counts by the total number of phonemes or words read, respectively, to account for incomplete passage completion; Visualized significance used t-tests (i.e., unadjusted for covariates), *p<.05, **p<.01, ***p<.001, NS=non-signficant; lvPPA = logopenic variant primary progressive aphasia (PPA); nfvPPA = nonfluent variant PPA;

**Table 3.** Group differences in phoneme- and word-level errors including pairwise comparisons.

| **Feature** | **ANCOVA *F* (df)** | ***p*** | **Pairwise Comparison** | **Mean Difference** | ***p* (FDR-corrected)** |
| --- | --- | --- | --- | --- | --- |
| *Phoneme insertion*<br>(per phoneme) | 5.37 (2, 94) | .006 | Control - lvPPA | -.007 | .511 |
|  |  |  | Control - nfvPPA | -.026 | .023 |
|  |  |  | lvPPA - nfvPPA | -.019 | .023 |
| *Phoneme replacement*<br>(per phoneme) | 14.83 (2, 94) | <.001 | Control - lvPPA | -.008 | .084 |
|  |  |  | Control - nfvPPA | -.021 | <.001 |
|  |  |  | lvPPA - nfvPPA | -.012 | .001 |
| *Phoneme repetition*<br>(per phoneme) | 6.57 (2, 94) | .002 | Control - lvPPA | -.002 | .524 |
|  |  |  | Control - nfvPPA | -.009 | .010 |
|  |  |  | lvPPA - nfvPPA | -.007 | .010 |
| *Phoneme deletion*<br>(per phoneme) | 19.35 (2, 94) | <.001 | Control - lvPPA | -.006 | .781 |
|  |  |  | Control - nfvPPA | -.085 | <.001 |
|  |  |  | lvPPA - nfvPPA | -.079 | <.001 |
| *Phoneme prolongation*<br>(per phoneme) | 5.86 (2, 94) | .004 | Control - lvPPA | -.001 | .914 |
|  |  |  | Control - nfvPPA | -.027 | .024 |
|  |  |  | lvPPA - nfvPPA | -.026 | .013 |
| *Word insertion*<br>(per word) | 6.32 (2, 94) | .003 | Control - lvPPA | -.015 | .313 |
|  |  |  | Control - nfvPPA | -.042 | .008 |
|  |  |  | lvPPA - nfvPPA | -.027 | .023 |
| *Word replacement*<br>(per word) | 10.74 (2, 94) | <.001 | Control - lvPPA | .003 | .937 |
|  |  |  | Control - nfvPPA | -.096 | .003 |
|  |  |  | lvPPA - nfvPPA | -.099 | <.001 |
| *Word deletion*<br>(per word) | 8.13 (2, 94) | .001 | Control - lvPPA | .016 | .392 |
|  |  |  | Control - nfvPPA | -.035 | .055 |
|  |  |  | lvPPA - nfvPPA | -.050 | .001 |

Note. lvPPA = logopenic Primary Progressive Aphasia (PPA), nfvPPA = non-fluent PPA; ANCOVA = Analysis of Covariance; df = degrees of freedom; FDR = False Discovery Rate

### Validation with expert-rated Motor Speech Evaluation

The relationships between automated speech features and SLP MSE ratings within the nfvPPA group are visualized in Figure 3, which includes a correlation matrix, scatterplots, and histograms of all variables. Across the eight speech error (dysfluency) features, moderate positive correlations were observed with the global MSE score (sum of AOS and dysarthria ratings), ranging from *r* = .30 to .58 (p<.001-.064).

**Figure 3.**
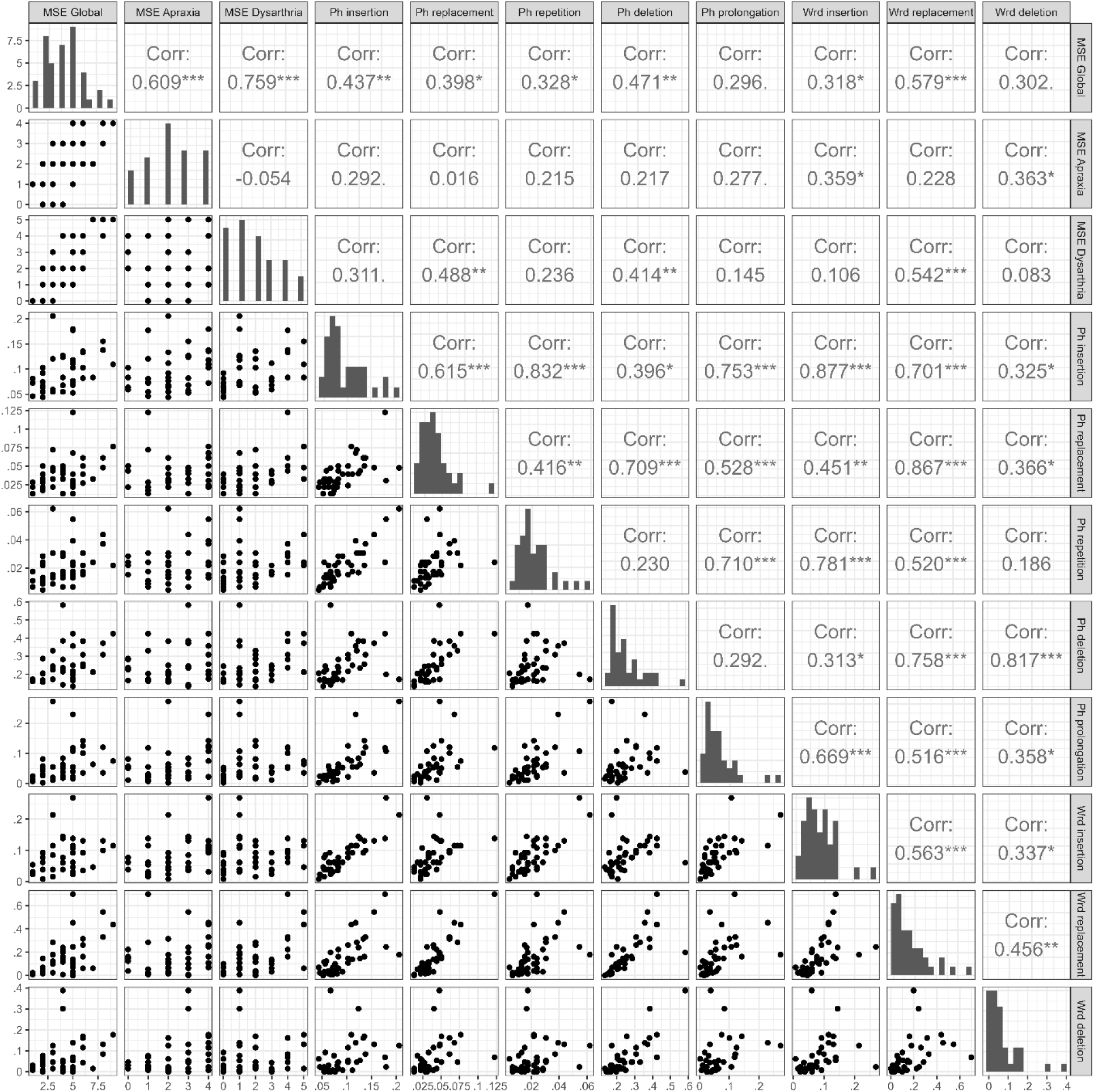
Scatterplot (lower triangle) and correlation matrix (upper triangle) of the three Motor Speech Evaluation (MSE) scores and the eight speech error (dysfluency) metrics, including histograms (diagonal);.p < .1, *p < .05,**p < .01, ***p < .001

### Machine learning classification

Figure 4 displays receiver operating characteristic (ROC) curves for classifying individuals with nfvPPA and lvPPA using automated speech error (dysfluency) features. A random forest machine learning model trained on the eight extracted features achieved an AUC of .806 [95% CI: .636–.976], indicating good classification performance based on speech errors alone. When age, education, and disease severity were added, model performance remained similar with an AUC of .805 [.631–.978]. An XGBoost classifier yielded similar results, with an AUC of .776 [.590–.961] using the speech features alone, and an improved AUC of .831 [.673–.989] when demographic and clinical variables were included. These converging results across two machine learning classifiers support the robustness of the automated features in distinguishing between PPA variants.

**Figure 4.**
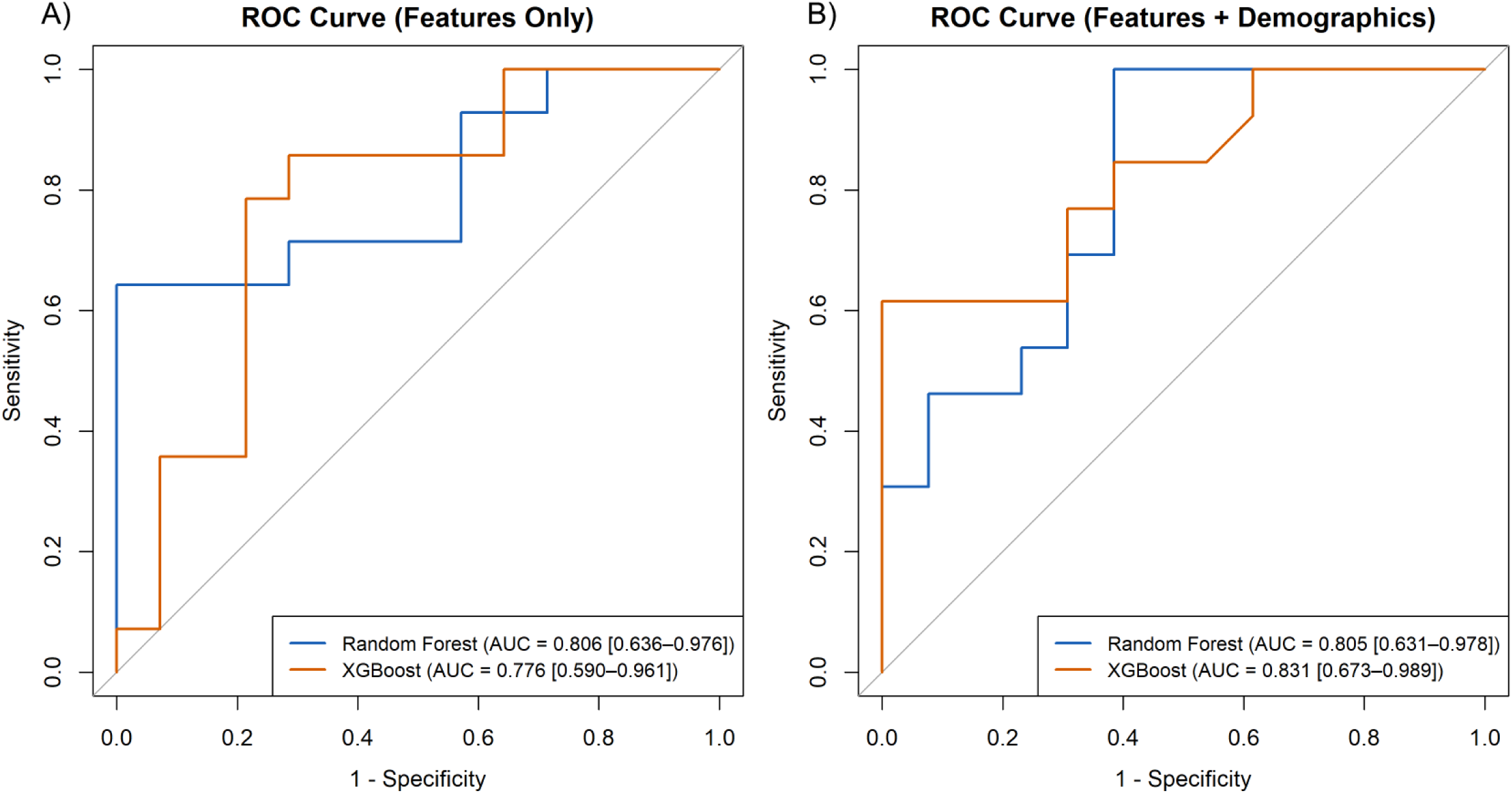
Receiver operating characteristic (ROC) curves for classifying nfvPPA versus lvPPA

## Discussion

This study demonstrated that AI-based automated analysis of speech errors (dysfluencies) extracted using the novel SSDM-L system, can detect PPA from controls and differentiate between individuals with nfvPPA and lvPPA. Using voice recordings from a brief reading task, the system captured dysfluencies that reflected known differences in speech production error profiles between variants. All features showed group differences, with the nfvPPA group making the most errors, followed by lvPPA, with minimal errors in cognitively healthy controls. These automated features also showed correlations with expert SLP-rated scores of motor speech impairment, confirming that an AI-based classification of speech dysfluencies aligns with clinical judgment as a preliminary proof-of-concept analysis. Finally, classification performance was good based on features alone, and further improved when demographic and disease severity variables were added to the model. Taken together, these findings suggest that the SSDM-L AI system provides a promising solution to automatically detect speech errors in connected speech.

Our findings provide a crucial advance to a growing body of work using automated speech analysis to study neurodegenerative disease, including PPA.^32,39–44^ A handful of studies have successfully applied natural language processing and audio signal processing techniques to characterize broad linguistic and/or acoustic patterns in PPA, such as reduced lexical diversity, simplified syntax, and semantic impairments.^32,39,41^ Our findings address a key limitation of previous off-the-shelf ASR tools, which prioritize fluent, intelligible output and systematically normalize dysfluencies, making them ill-suited for detecting speech errors that are crucial to the diagnosis and monitoring of PPA, and other neurological diseases. Dysfluency detection remains one of the most persistent challenges in ASR, particularly in clinical populations where speech is degraded or non-standard.^6^ Our study provides evidence that the SSDM-L system, combining phoneme-level alignment with rule-based detection, enables the capture of clinically meaningful speech disruptions automatically and efficiently. As automated speech tools gain traction in both clinical and research settings, developing systems that preserve diagnostically informative signals (rather than correcting them away) will be critical for advancing their clinical utility, particularly in disorders such as PPA where speech production deficits are a defining feature.

The machine learning classification results further support the potential of automated speech error (dysfluency) analysis for assisting in differential diagnosis of PPA. Using SSDM-L features, the models achieved good discrimination between nfvPPA and lvPPA, demonstrating that fine-grained phoneme- and word-level errors carry meaningful diagnostic signal. Models remained similar or only slightly improved after adding demographic and clinical variables, likely since groups were matched on these variables. These findings are consistent with prior studies applying machine learning to speech and language features in PPA and other neurodegenerative syndromes, which have similarly shown promising accuracy in distinguishing subtypes.^43,45^

The prominence of motor speech impairments in individuals with nfvPPA has been well-documented.^9^ AOS is one of the two core diagnostic criteria for nfvPPA and is frequently the earliest and most prominent symptom;^8^ dysarthria is also often observed in individuals with nfvPPA.^46,47^ Consistently, our participants with nfvPPA showed significantly higher scores in AOS and dysarthria severity than those with lvPPA on the motor speech evaluation carried out by SLP experts. Determining the presence of motor speech impairments typically requires expert perceptual judgment by an SLP—a process that is time-consuming, resource-intensive, and often limited by the scarcity of clinicians with specialized expertise in dementia clinics.^48,49^ The automated speech features identified in this study were consistently correlated with expert SLP-rated global MSE scores in the nfvPPA group, supporting the interpretation that higher feature counts are at least partially related to underlying motor speech impairments. These findings reinforce the construct validity of the SSDM-L-derived metrics and suggest that automated analysis may offer a scalable complement to clinician-based assessments of speech errors in PPA. Further studies are necessary to determine whether the AI system can match the precision of clinicians in identifying specific error types (e.g., differentiating dysarthria and AOS) in individual cases.

A core strength of our approach is that it is grounded in expert clinical knowledge, incorporating input from trained SLPs and aligning with real-world observations of speech behavior in PPA. Using the Grandfather Passage allowed us to control lexical content and isolate dysfluency patterns, avoiding confounds related to language formulation and vocabulary access, while the use of forced alignment enabled precise detection of phonemic deviations that reflect underlying motor or phonological impairments. Nonetheless, several limitations should be noted. Although our sample sizes are comparable to those in other studies of PPA,^39,43,50,51^ they remain relatively small for achieving optimal statistical power and broad generalizability. Nonetheless, the consistent group differences observed across all features suggest that the automated measures are robust and sensitive to meaningful speech impairments, even in a modestly-sized sample. In addition, all participants were native English speakers, which may restrict the applicability of the SSDM-L system to individuals with different linguistic backgrounds. Additionally, the current analysis focused on a single speech elicitation task. Although the Grandfather Passage offers important advantages for controlled error analysis, future research should examine the generalizability of these findings to spontaneous speech tasks such as picture description^52^ or personal narratives.^53^ Finally, a limitation of the AI based system is that similar coding conflicts that exist within human-expert annotators can manifest within AI-based automated error analysis. For example, a single phoneme replacement error can equivalently be represented as one deletion followed by one insertion. While such distinctions can often be resolved by trained SLPs, current AI-based error analyzers (including SSDM-L) are not sufficiently precise to consistently disambiguate these mechanistically equivalent error modes. Future refinements of the AI system that integrate more SLP-guided error classifications are likely to further improve both the accuracy and consistency of automated error analysis beyond what we have demonstrated in the present study.

Our study provides strong evidence of the potential of AI methods to transform how speech errors are identified and monitored in neurodegenerative diseases, particularly in under-resourced settings where expertise in speech-language pathology is limited or unavailable. The integration of engineered speech analytics into routine care could bring the field closer to more equitable, efficient, and precise models of diagnosis and disease tracking in neurodegenerative conditions.

## Acknowledgments

This work was supported by the National Institutes of Health (NIH) under the following grants: R01NS050915, K24DC015544, R01AG075775, P01AG019724 (M.L. Gorno-Tempini) and R00AG066934, RF1AG091509 (J.M.J. Vonk). We are grateful to our participants and their families for participating in this research.

## Notes

### Competing Interest Statement

The authors have declared no competing interest.

### Author Declarations

All participants or caregivers provided informed consent following procedures aligned with the Declaration of Helsinki, and the study was approved by the UCSF Institutional Review Board (IRB # 10-03946).

